## Supplementary Material for "Active remodeling of the chromatin landscape directs extravillous trophoblast cell lineage development"

**This PDF file includes:**

Supplementary Figures 1-25
Supplementary References

**Other supplementary materials for this manuscript include the following:**

Supplementary Tables 1-21 (separate excel file)

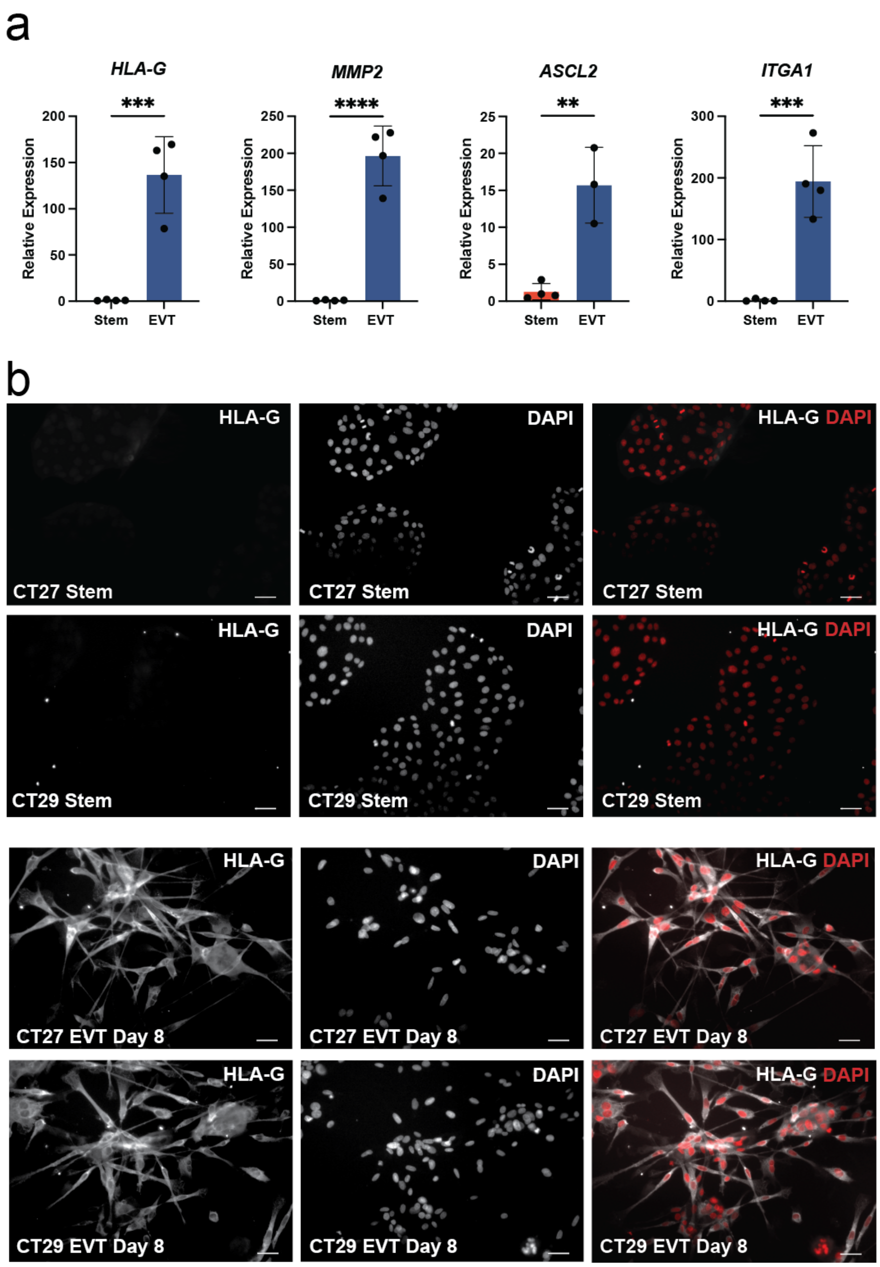

**Supplementary Figure 1. Expression of EVT cell state-specific markers including HLA-G.**

**a)** Relative expression of *HLA-G*, *MMP2*, *ASCL2*, and *ITGA1* measured by RT-qPCR in stem state and following EVT cell differentiation (n=4 per group) (**$p<0.01$, ***$p<0.001$, ****$p<0.0001$). Graphs depict mean ± standard deviation. **b)** EVT cell differentiation is accompanied by cell elongation and upregulation of HLA-G (white) in CT27 and CT29 cell lines. DAPI stains cell nuclei (white or red as labeled). Scale bars represent 100 μm.

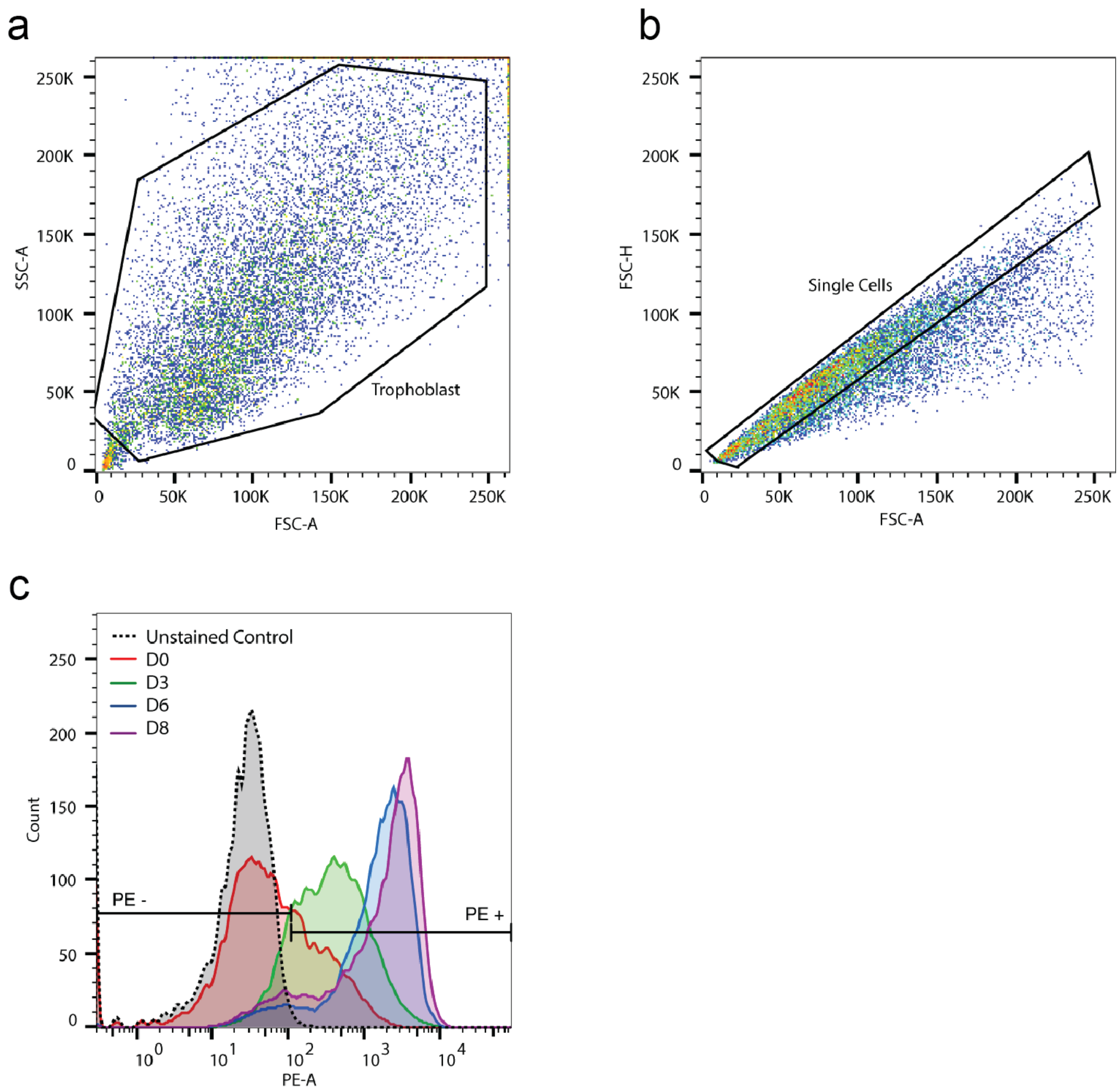

**Supplementary Figure 2. Gating strategy for flow cytometric measurements of HLA-G.** Representative plots depicting the gating strategy used to select for **a)** trophoblast cells (x-axis, forward scatter area, **FSC-A**; y-axis, side scatter area, SSC-A), followed by **b)** single cells (x-axis FSC-A; y-axis, forward scatter height, **FSC-H**), and then **c)** HLA-G positive cells (Phycoerythrin, **PE**+) depicted by black gates (x-axis, phycoerythrin area, PE-A; y-axis Count). Unstained control (black dotted line), stem state (D0 of EVT cell differentiation; red), day 3 (green), day 6 (blue), and day 8 (purple) of EVT cell differentiation are shown.

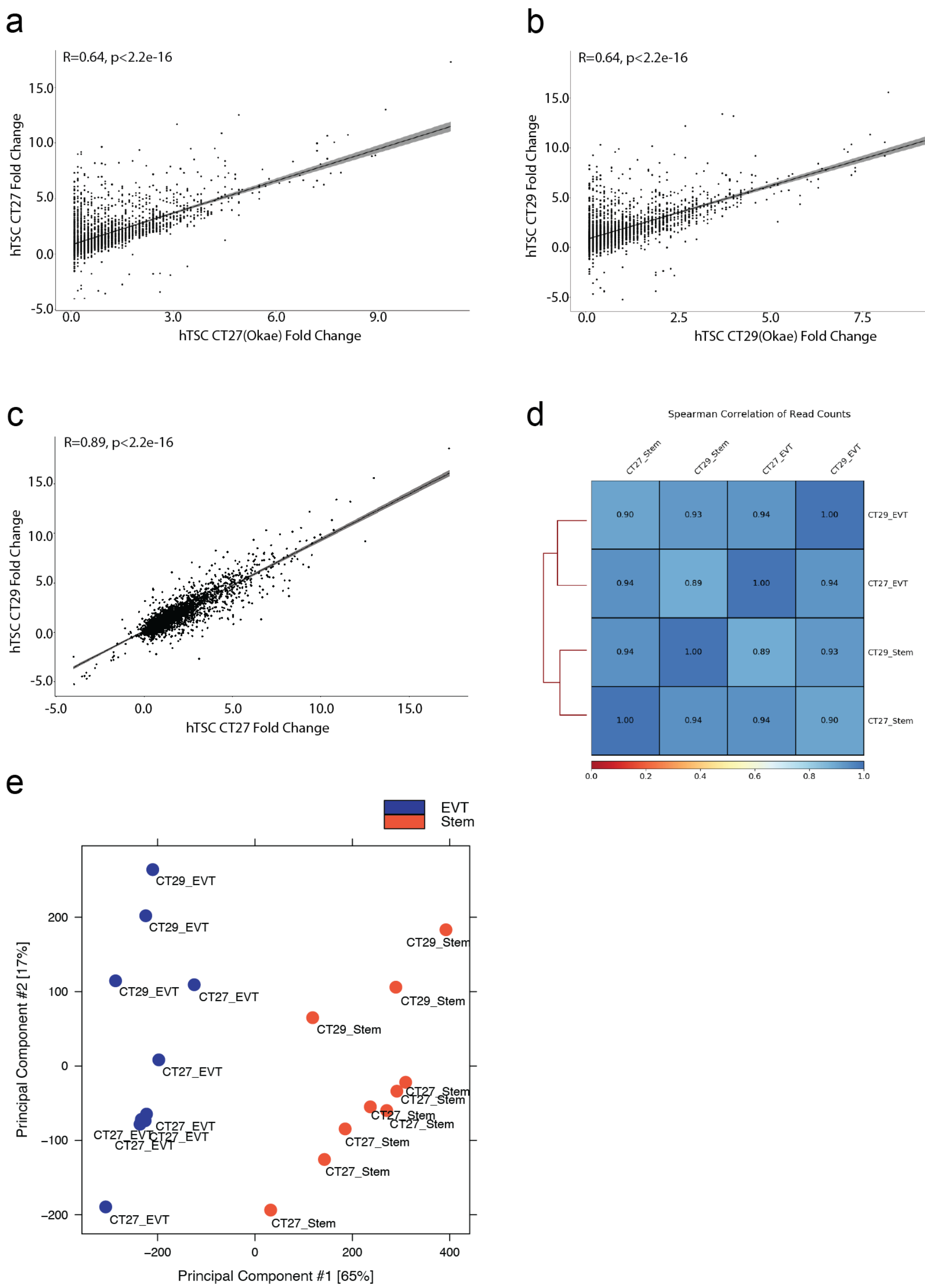

**Supplementary Figure 3. Validation of transcriptomes across human TS cell donor lines and datasets.** Scatter plots comparing log2 fold change from differential expression analysis of EVT cell transcriptomes relative to stem state transcriptomes in the CT27 (**a**) and CT29 (**b**) TS cell lines measured by RNA-Seq in this report (y-axis) and the Okae et al. (2018) report[1] (x-axis). Pearson correlations were calculated to estimate strength of the linear association. **c)** Scatter plots comparing log2 fold change from differential expression analysis of EVT cell transcriptomes relative to stem state transcriptomes for CT27 (x-axis) and CT29 (y-axis) TS cell lines generated from RNA-Seq and presented in this report. Pearson correlation was calculated to estimate strength of the linear association. **d)** Spearman correlation heatmap using affinity (read count) data for EVT and Stem cell ATAC-Seq libraries for CT27 and CT29 TS cell lines. **e)** Principal component analysis (**PCA**) plot depicting EVT (blue) and Stem (red) cell data of counts per million mapped reads for independent ATAC-Seq libraries.

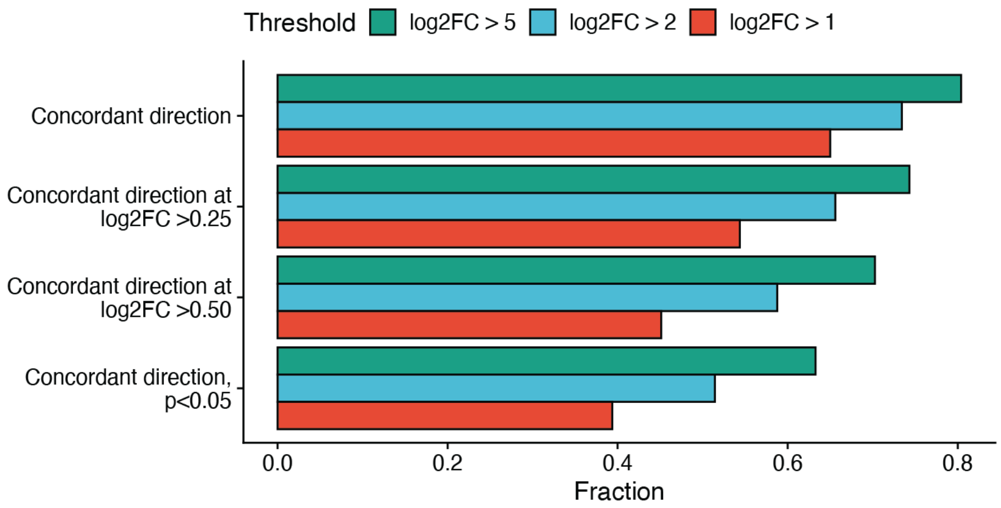

**Supplementary Figure 4. Validation of gene expression patterns in EVT cells between *in vitro* and *in vivo* data sets.** Bar graphs depicting concordance of differentially expressed genes detected in human TS cells (*in vitro*) compared to single-cell RNA expression data in first trimester placenta samples (*in vivo*). Concordance (Fraction, x-axis) is estimated for genes differently expressed in EVT cells compared to stem state cells at log2 fold-change > 5 (green bars), log2 fold-change > 2 (blue bars) and log2 fold-change > 1 (red bars) based on different threshold for differential expression in the *in vivo* data sets (y axis).

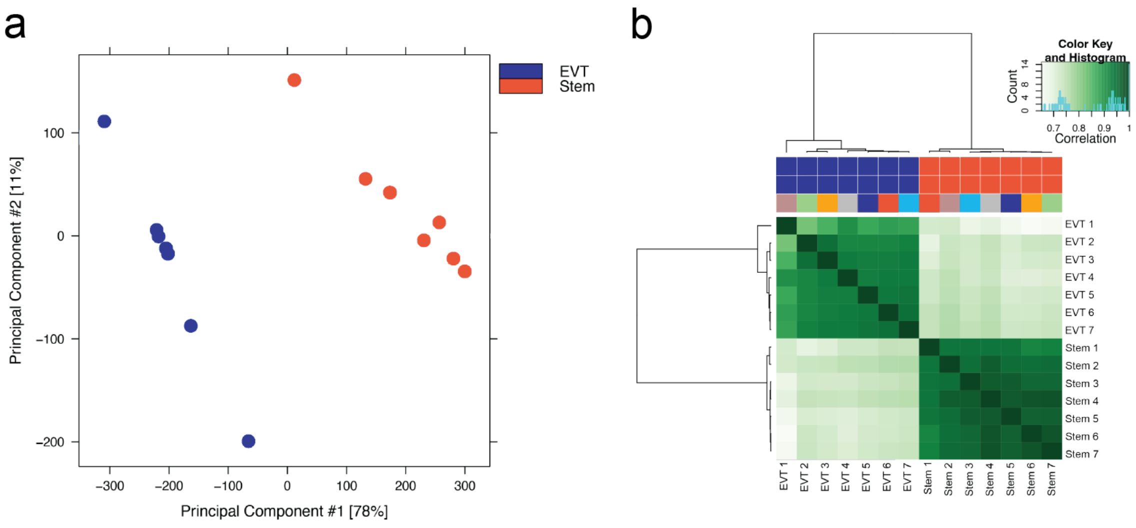

**Supplementary Figure 5. Stem and EVT cell-differentiated human TS cells segregation based on chromatin accessibility. a)** Principal component analysis (**PCA**) plot depicting EVT (blue) and Stem (orange) cell data of normalized read counts for independent ATAC-Seq libraries for all regions tested in the CT27 cell line. **b)** Correlation heatmap using affinity (read count) data for EVT (blue) and Stem (orange) cell ATAC-Seq libraries generated from the CT27 cell line.

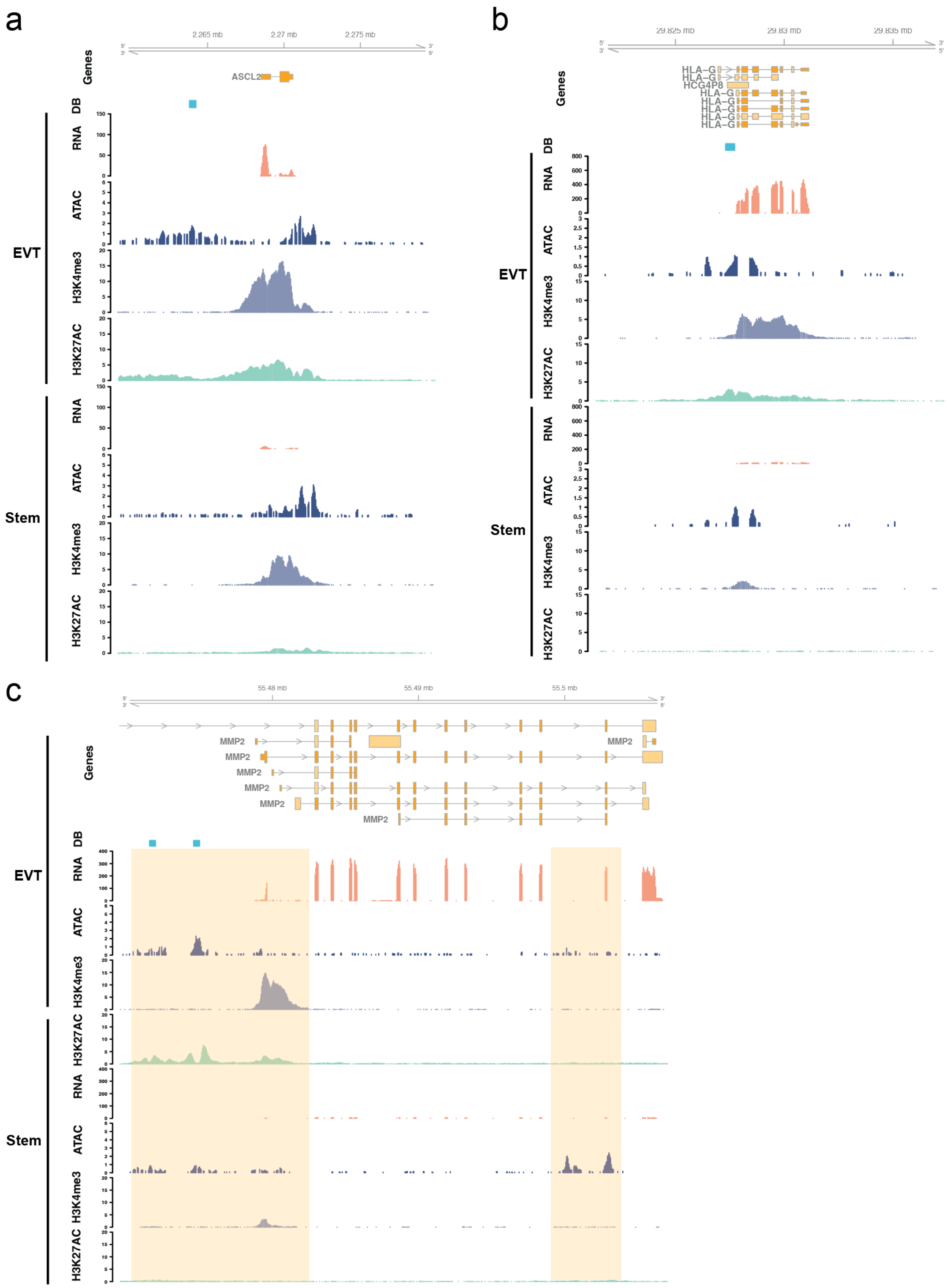

**Supplementary Figure 6. Regulatory landscape near EVT cell-specific gene regions in CT27 cells. a-c)** RNA-Seq, ATAC-Seq, histone 3 lysine 4 trimethylation (**H3K4me3**) and histone 3 lysine 27 acetylation (**H3K27ac**) ChIP-Seq assessments performed in EVT cells (top panel), and stem state cells (bottom panel) are shown for regions surrounding *ASCL2* **(a)**, *HLA-G* **(b)**, and *MMP2* **(c)** in CT27 cells. Differentially bound (**DB**) regions are specific to the EVT cell state and are shown in blue rectangles. RNA-Seq (transcripts per million, y-axis), ATAC-Seq (counts per million mapped reads, y-axis), H3K4me3, and H3K27ac ChIP-Seq (counts per million mapped reads, y-axis) datasets are shown in individual tracks. Key regulatory regions are highlighted in yellow.

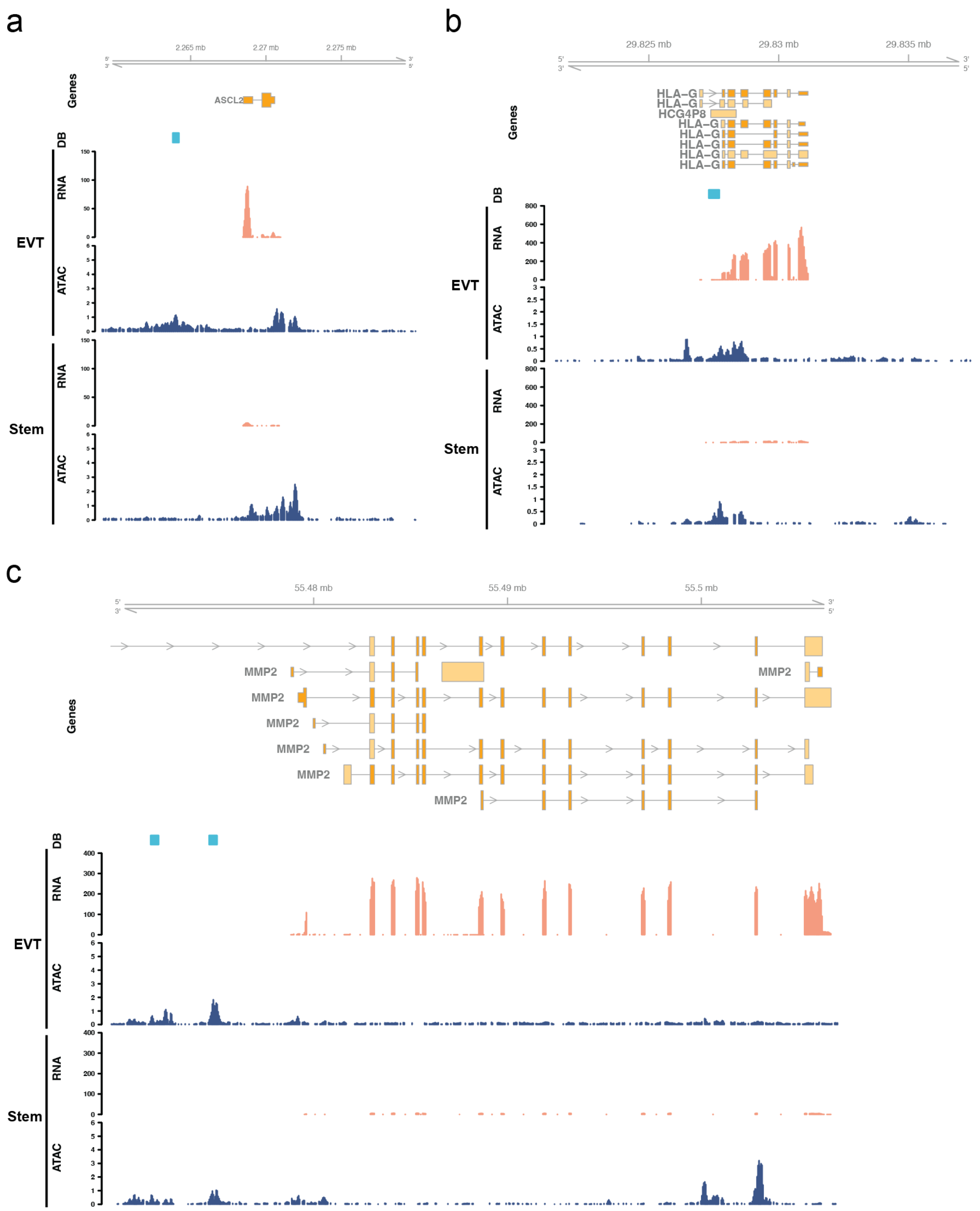

**Supplementary Figure 7. Regulatory landscape near EVT-specific gene regions in CT29 cells. a-c)** RNA-Seq and ATAC-Seq assessments performed in EVT cells (top panel), and stem state cells (bottom panel) are shown for regions surrounding *ASCL2* **(a)**, *HLA-G* **(b)**, and *MMP2* **(c)** in CT29 cells. Differentially bound (**DB**) regions are specific to the EVT cell state and are shown in blue rectangles. RNA-Seq (transcripts per million, y-axis) and ATAC-Seq (counts per million mapped reads, y-axis) datasets are shown in individual tracks.

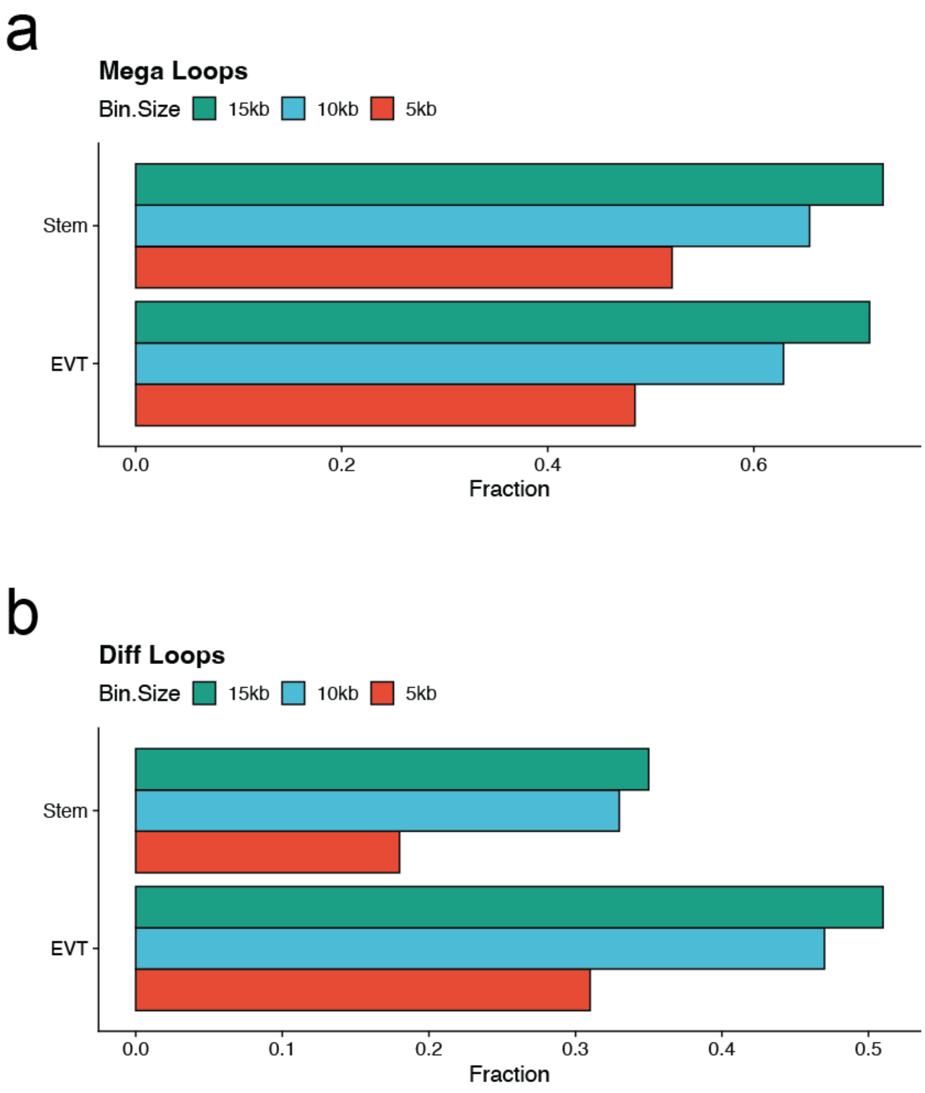

**Supplementary Figure 8. Validation of long-range chromatin interactions in stem state and EVT differentiated cells. a-b)** Bar graphs depicting the fraction (x-axis) of all Hi-C chromatin loops detected (**a**; Mega) or those unique (**b**; Diff) to either stem or EVT cell states in CT27 cell that are also identified in the CT29 cell line within 5kb (red), 10kb (blue) or 15kb (green) of both anchors.

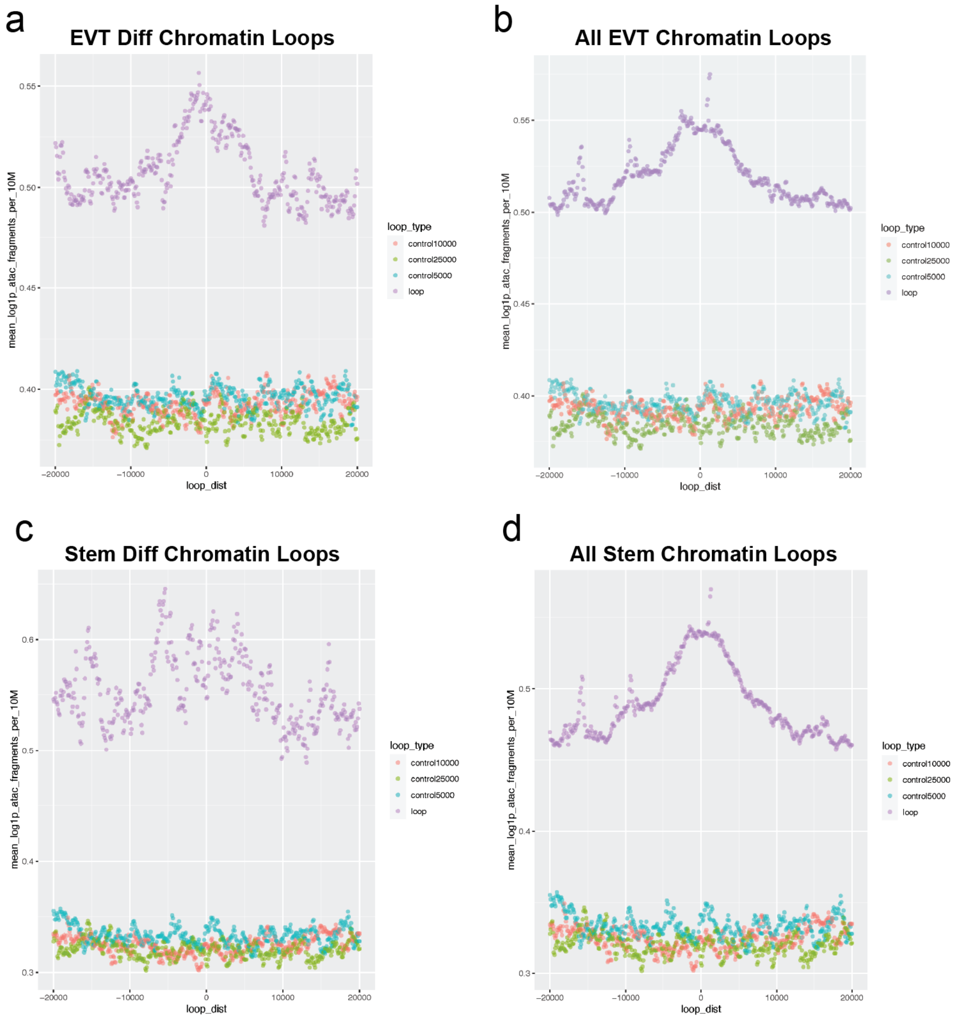

**Supplementary Figure 9. Chromatin accessibility at loop anchors.** Density plots showing distribution of open chromatin relative to the center of loop anchors (bp, x-axis). Normalized bin densities for every 10 million total reads (y-axis) as measured by ATAC-seq are shown for chromatin loops called in **a**) EVT Diff loops, **b**) All EVT loops, **c**) Stem Diff loops and **d**) All Stem loops as shown in purple. Other regions below the peak signals indicate the results from 1000 randomly selected control sets that do not intersect any of the loops with region sizes of 5 kb (blue), 10 kb (red) and 25 kb (green).

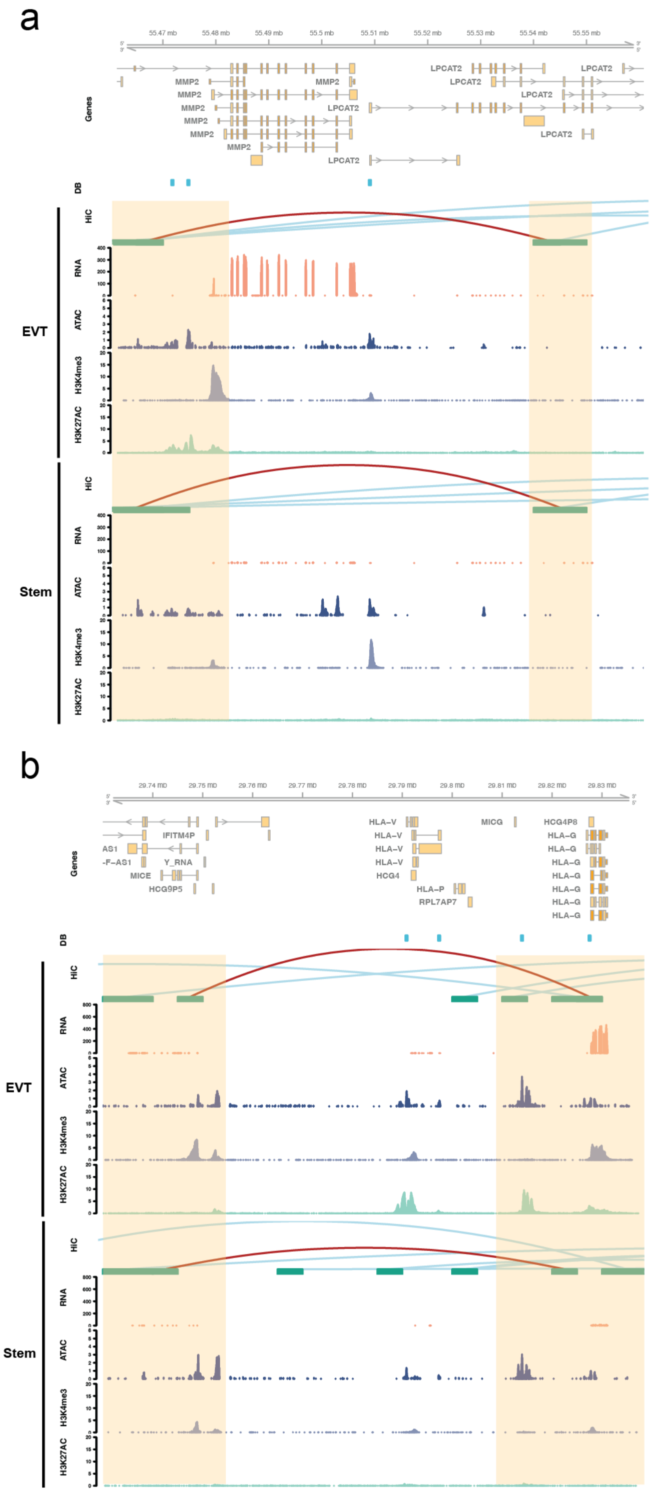

**Supplementary Figure 10. Long-range chromatin interactions near *MMP2* and *HLA-G* in CT27 cells. a, b)** Hi-C chromatin capture, RNA-Seq, ATAC-Seq, and H3K4me3 and H3K27ac ChIP-Seq assessments performed in EVT cells (top panel), and stem state cells (bottom panel)

are shown for regions surrounding *MMP2* **(a)** and *HLA-G* **(b)** in CT27 cells. Differentially bound (**DB**) regions are specific to the EVT cell state and are shown in blue rectangles. Hi-C loops (red, both loop anchors in view; blue, loop anchors out of view), RNA-Seq (transcripts per million, y-axis), ATAC-Seq (counts per million mapped reads, y-axis), H3K4me3, and H3K27ac ChIP-Seq (counts per million mapped reads, y-axis) datasets are shown in individual tracks. Key regulatory regions are highlighted in yellow.

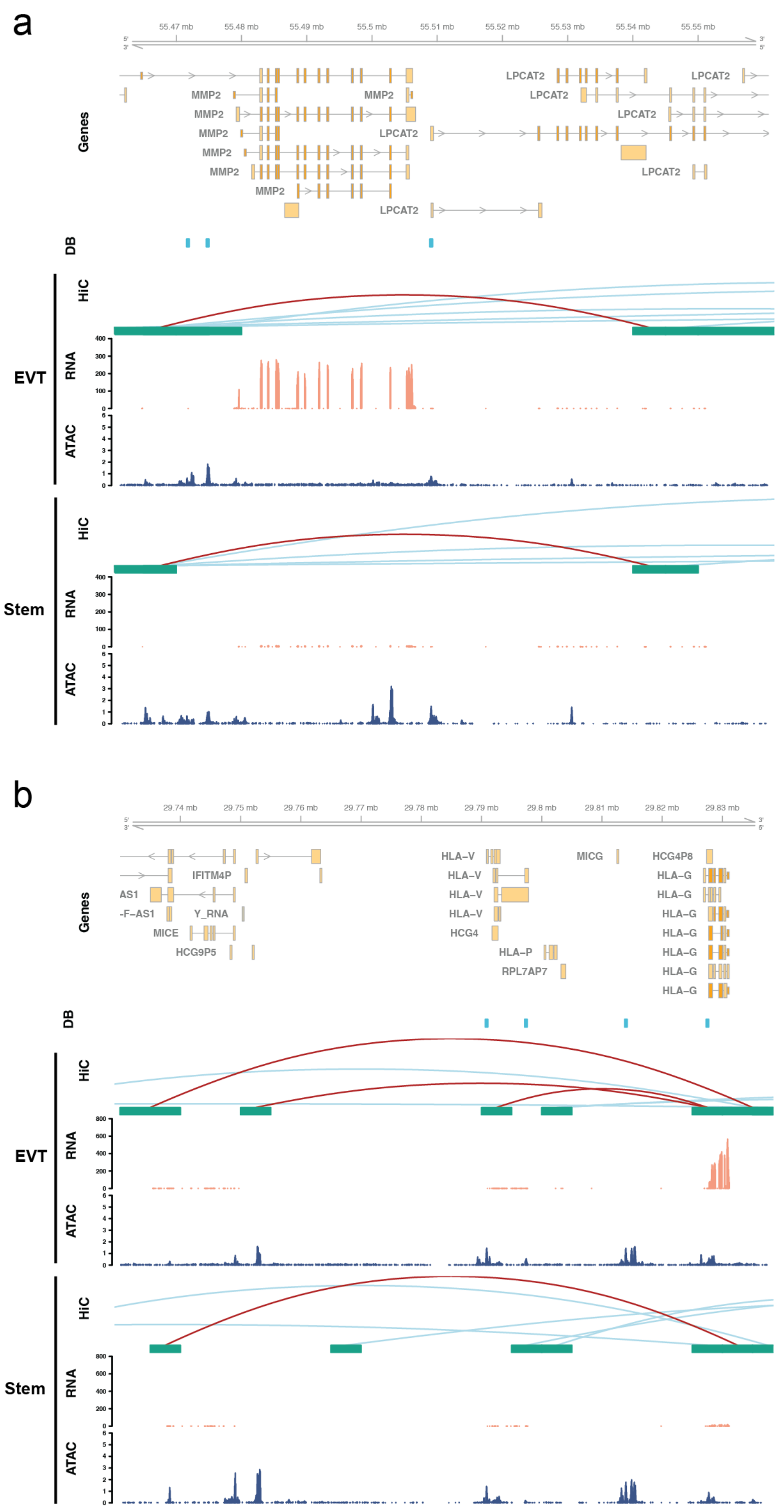

**Supplementary Figure 11. Long-range chromatin interactions near *MMP2* and *HLA-G* in CT29 cells. a, b)** Hi-C chromatin capture, RNA-Seq, and ATAC-Seq assessments performed in EVT cells (top panel), and stem state cells (bottom panel) are shown for regions surrounding

*MMP2* **(a)** and *HLA-G* **(b)** in CT29 cells. Differentially bound (**DB**) regions are specific to the EVT cell state and are shown in blue rectangles. Hi-C loops (red, both loop anchors in view; blue, loop anchors out of view), RNA-Seq (transcripts per million, y-axis), and ATAC-Seq (counts per million mapped reads, y-axis) datasets are shown in individual tracks.

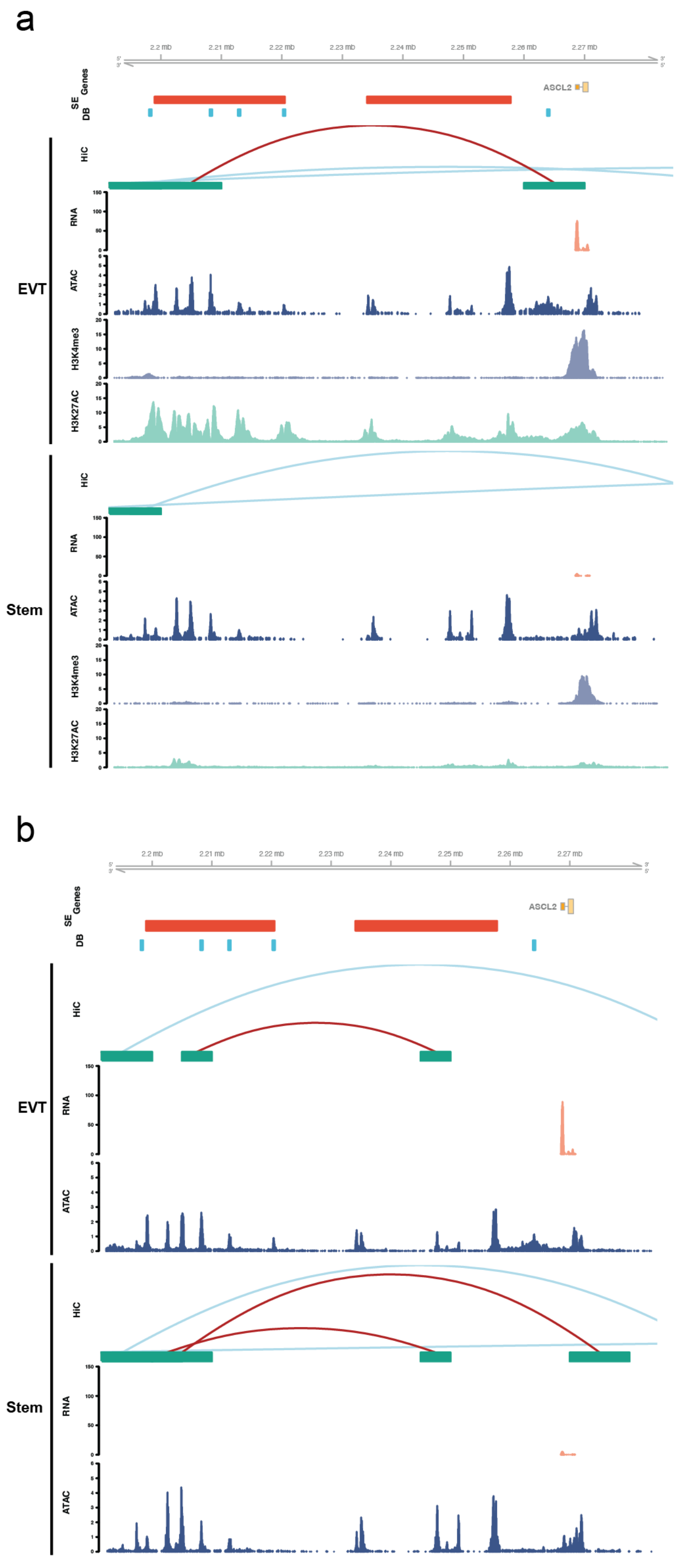

**Supplementary Figure 12. Long-range chromatin interactions defining a unique regulatory landscape near *ASCL2* in EVT cells. a)** Hi-C, RNA-Seq, ATAC-Seq, and

H3K4me3 and H3K27ac ChIP-Seq assessments performed in CT27 cells differentiated into EVT cells (top panel) or maintained in the stem state (bottom panel). Regulatory elements near *ASCL2* (yellow) are shown and include Hi-C loops (red, both loop anchors in view; blue, loop anchors out of view), RNA-Seq (transcripts per million, y-axis), ATAC-Seq (counts per million mapped reads, y-axis), H3K4me3, and H3K27ac ChIP-Seq (counts per million mapped reads, y-axis). All datasets are shown in individual tracks. Super-enhancers (**SE**, red) and differentially bound regions (**DB**, blue) are specific to the EVT cell state. **b)** Hi-C, RNA-Seq, and ATAC-Seq assessments performed in CT29 cells differentiated into EVT cells (top panel) or maintained in the stem state (bottom panel). Regulatory elements near *ASCL2* (yellow) are shown and include Hi-C loops (red, both loop anchors in view; blue, loop anchors out of view), RNA-Seq (transcripts per million, y-axis), and ATAC-Seq (counts per million mapped reads, y-axis). All datasets are shown in individual tracks. Super-enhancers (**SE**, red) and differentially bound regions (**DB**, blue) are specific to the EVT cell state.

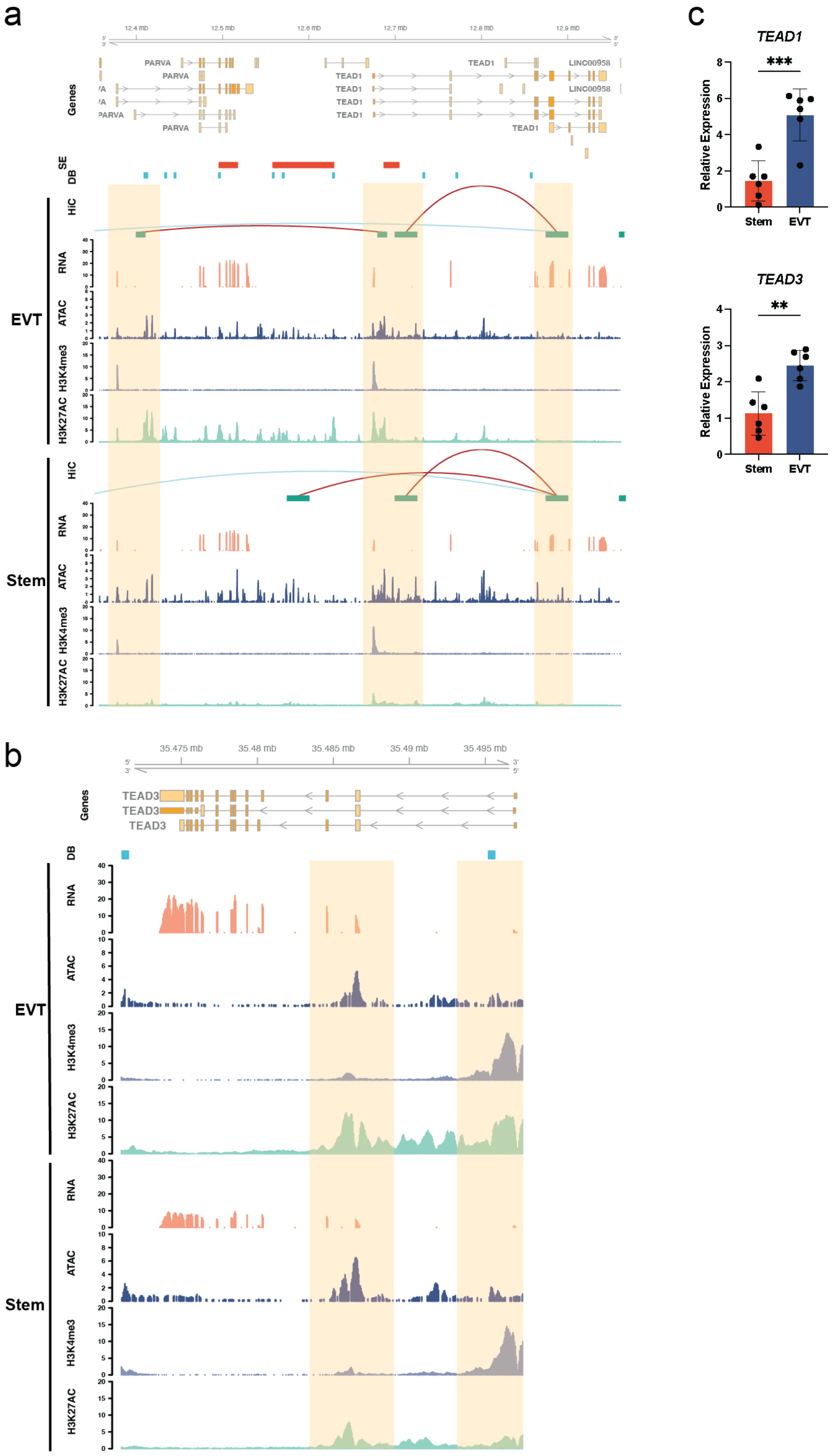

**Supplementary Figure 13. Long-range chromatin interactions near genes of the TEAD transcription factor family in CT27 cells.** Hi-C, RNA-Seq, ATAC-Seq, and H3K4me3 and H3K27ac ChIP-Seq assessments performed in CT27 cells differentiated into EVT cells (top panel) or maintained in the stem state (bottom panel). Regulatory elements near *TEAD1* (**a**) and *TEAD3* (**b**) are shown and include Hi-C loops (red, both loop anchors in view; blue, loop anchors out of view), RNA-Seq (transcripts per million, y-axis), ATAC-Seq (counts per million mapped reads, y-axis), H3K4me3, and H3K27ac ChIP-Seq (counts per million mapped reads, y-axis). All datasets are shown in individual tracks. Super-enhancers (**SE**, red) and differentially bound regions (**DB**, blue) are specific to the EVT cell state. Key regulatory regions are highlighted in yellow. **c)** The relative expression of *TEAD1*, and *TEAD3* in stem state (red; n=6 per group) and EVT cells (blue; n=6 per group) measured by RT-qPCR (**$p<0.01$ and ***$p<0.001$). Graphs depict mean ± SD.

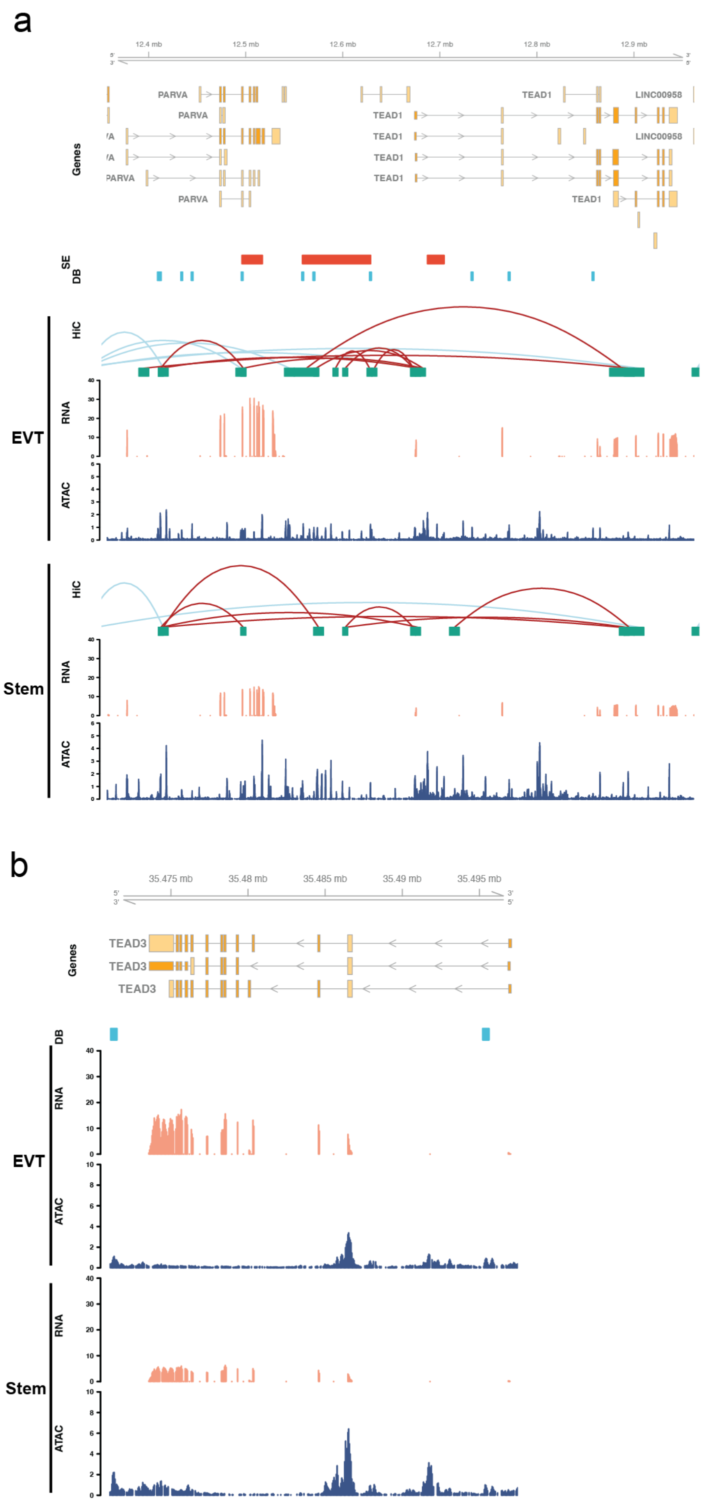

**Supplementary Figure 14. Long-range chromatin interactions near genes of the TEAD transcription factor family in CT29 cells.** Hi-C, RNA-Seq, and ATAC-Seq assessments performed in CT29 cells differentiated into EVT cells (top panel) or maintained in the stem state (bottom panel). Regulatory elements near *TEAD1* (**a**) and *TEAD3* (**b**) are shown and include Hi-C loops (red, both loop anchors in view; blue, loop anchors out of view), RNA-Seq (transcripts per million, y-axis), and ATAC-Seq (counts per million mapped reads, y-axis). All datasets are shown in individual tracks. Super-enhancers (**SE**, red) and differentially bound regions (**DB**, blue) are specific to the EVT cell state.

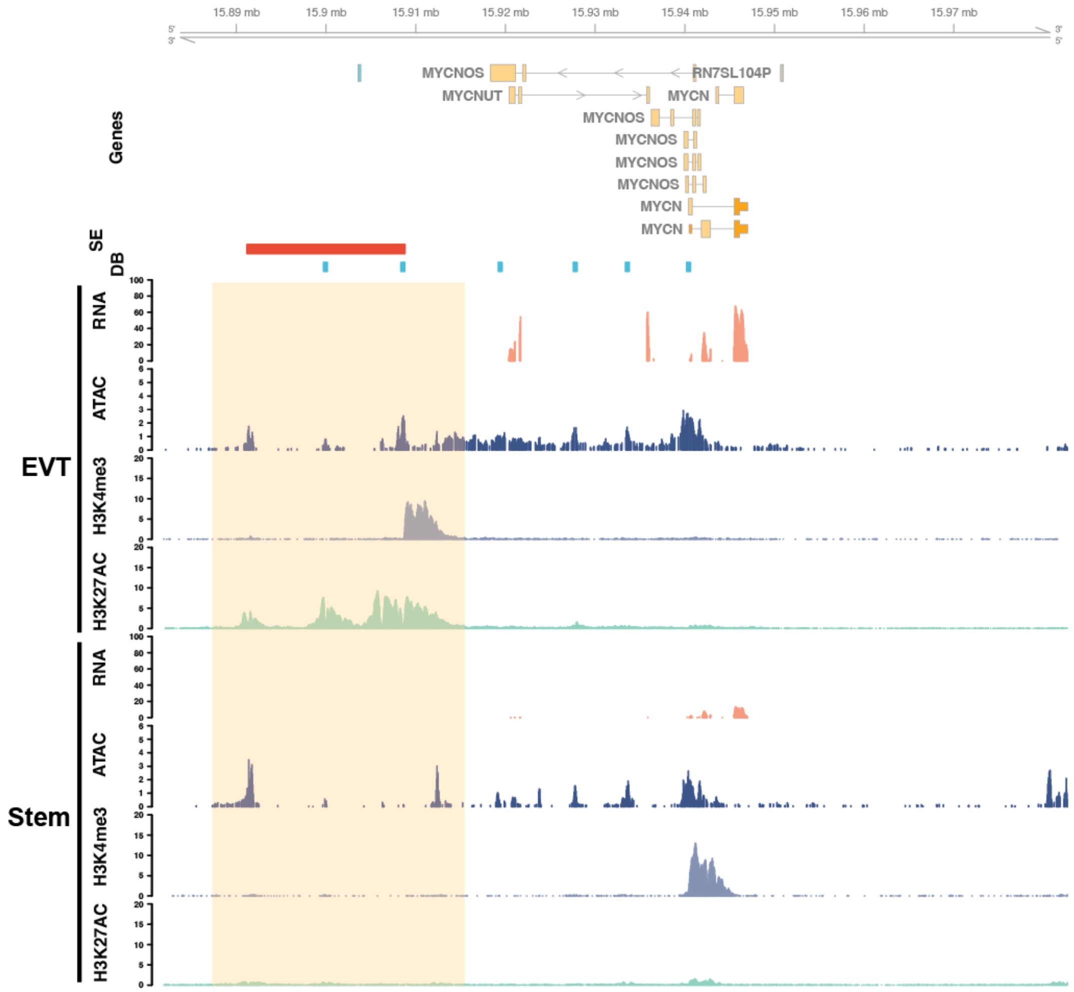

**Supplementary Figure 15. Long-range chromatin interactions near *MYCN* in EVT cell differentiated CT27 cells.** Hi-C, RNA-Seq, ATAC-Seq, and H3K4me3 and H3K27ac ChIP-Seq assessments performed in CT27 cells differentiated into EVT cells (top panel) or maintained in the stem state (bottom panel). Regulatory elements near *MYCN* are shown and include Hi-C loops (red, both loop anchors in view; blue, loop anchors out of view), RNA-Seq (transcripts per million, y-axis), ATAC-Seq (counts per million mapped reads, y-axis), H3K4me3, and H3K27ac ChIP-Seq (counts per million mapped reads, y-axis). All datasets are shown in individual tracks. Super-enhancers (**SE**, red) and differentially bound regions (**DB**, blue) are specific to the EVT cell state. Key regulatory regions are highlighted in yellow.

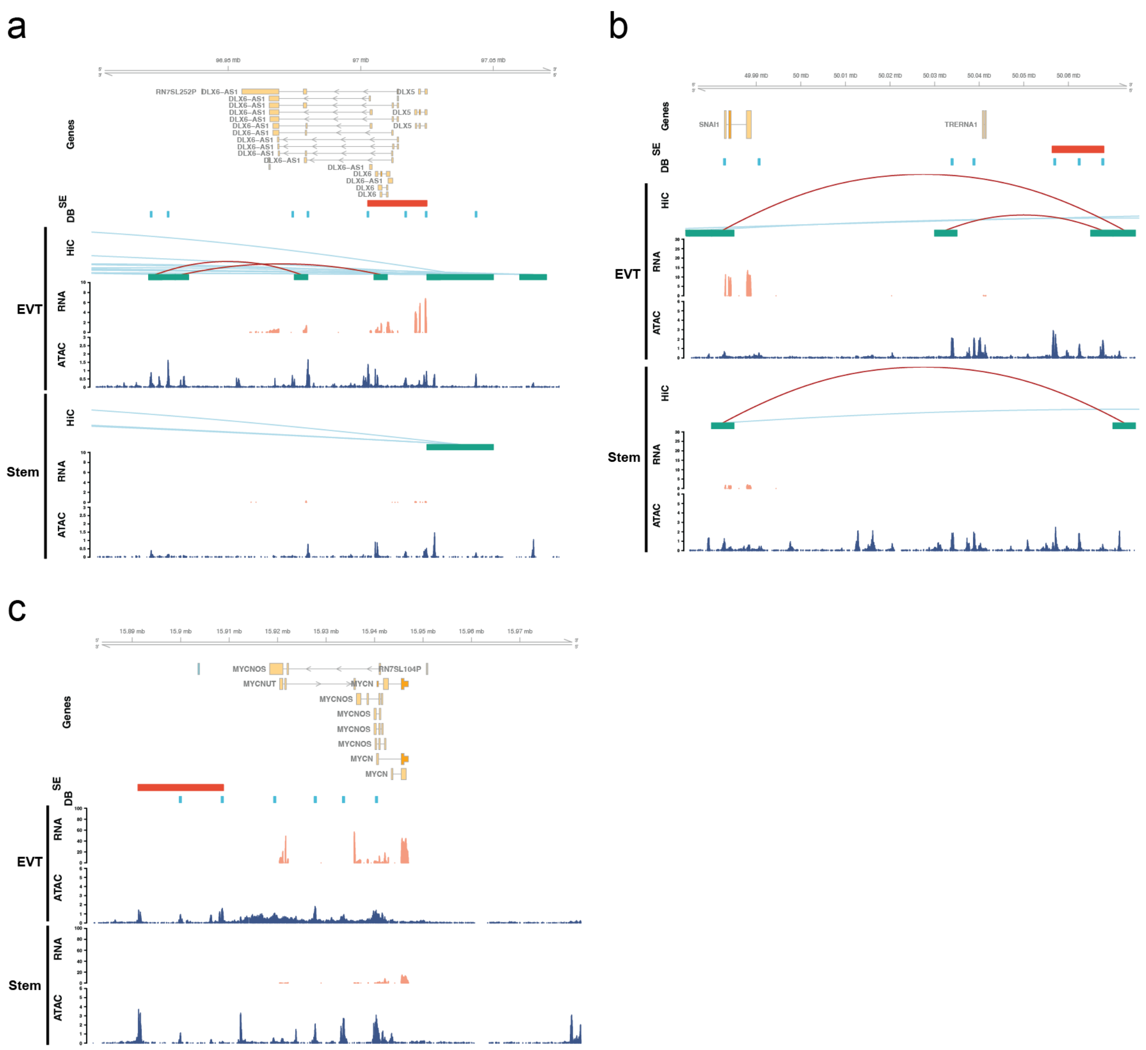

**Supplementary Figure 16. Long-range chromatin interactions for transcriptional regulators of EVT cell differentiation in CT29 cells.** Hi-C, RNA-Seq, and ATAC-Seq assessments performed in CT29 cells differentiated into EVT cells (top panel) or maintained in the stem state (bottom panel). Regulatory elements near *DLX5/DLX6* (**a**), *SNAI1* (**b**), and *MYCN* (**c**) are shown and include Hi-C loops (red, both loop anchors in view; blue, loop anchors out of view), RNA-Seq (transcripts per million, y-axis), and ATAC-Seq (counts per million mapped reads, y-axis). All datasets are shown in individual tracks. Super-enhancers (**SE**, red) and differentially bound regions (**DB**, blue) are specific to the EVT cell state.

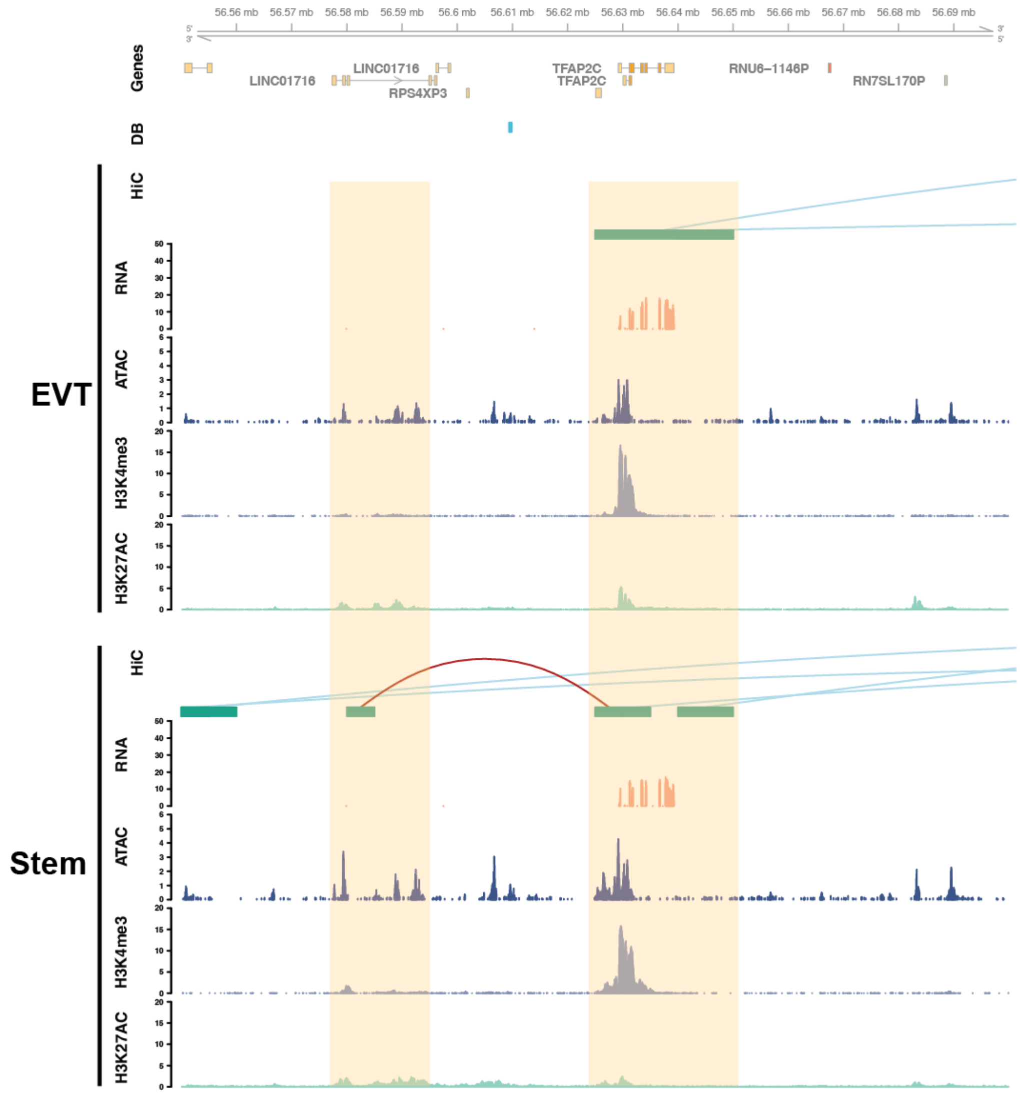

**Supplementary Figure 17. Regulatory landscape of *TFAP2C* in CT27 cells.** Hi-C, RNA-Seq, ATAC-Seq, and H3K4me3 and H3K27ac ChIP-Seq assessments performed in CT27 cells differentiated into EVT cells (top panel) or maintained in the stem state (bottom panel). Regulatory elements near *TFAP2C* are shown and include Hi-C loops (red, both loop anchors in view; blue, loop anchors out of view), RNA-Seq (transcripts per million, y-axis), ATAC-Seq (counts per million mapped reads, y-axis), K4me3, and K27ac ChIP-Seq (counts per million mapped reads, y-axis). All datasets are shown in individual tracks. Differentially bound regions (**DB**, blue) are specific to the EVT cell state. Key regulatory regions are highlighted in yellow.

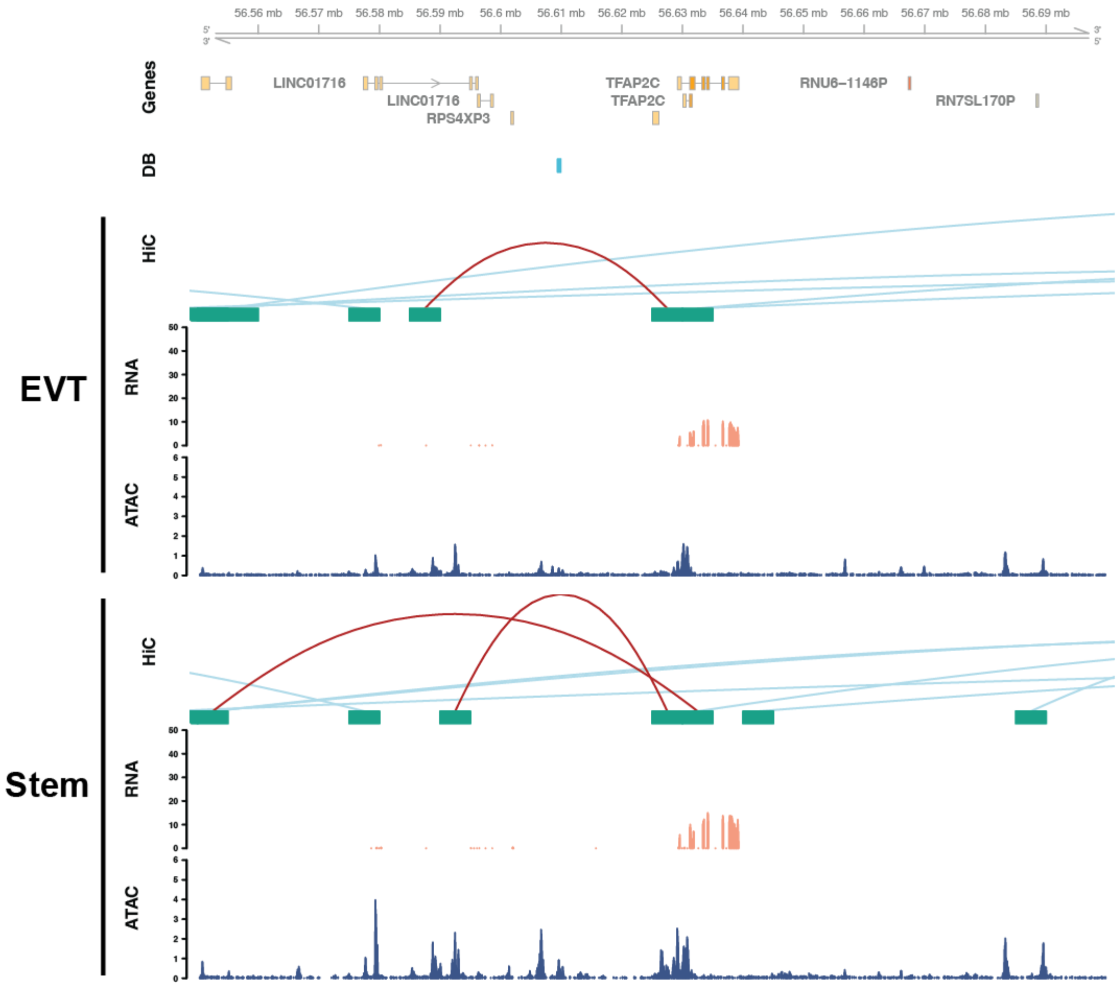

**Supplementary Figure 18. Long-range chromatin interactions near *TFAP2C* in CT29 cells.** Hi-C, RNA-Seq, and ATAC-Seq assessments performed in CT29 cells differentiated into EVT cells (top panel) or maintained in the stem state (bottom panel). Regulatory elements near *TFAP2C* are shown and include Hi-C loops (red, both loop anchors in view; blue, loop anchors out of view), RNA-Seq (transcripts per million, y-axis), and ATAC-Seq (counts per million mapped reads, y-axis). All datasets are shown in individual tracks. Differentially bound regions (**DB**, blue) are specific to the EVT cell state.

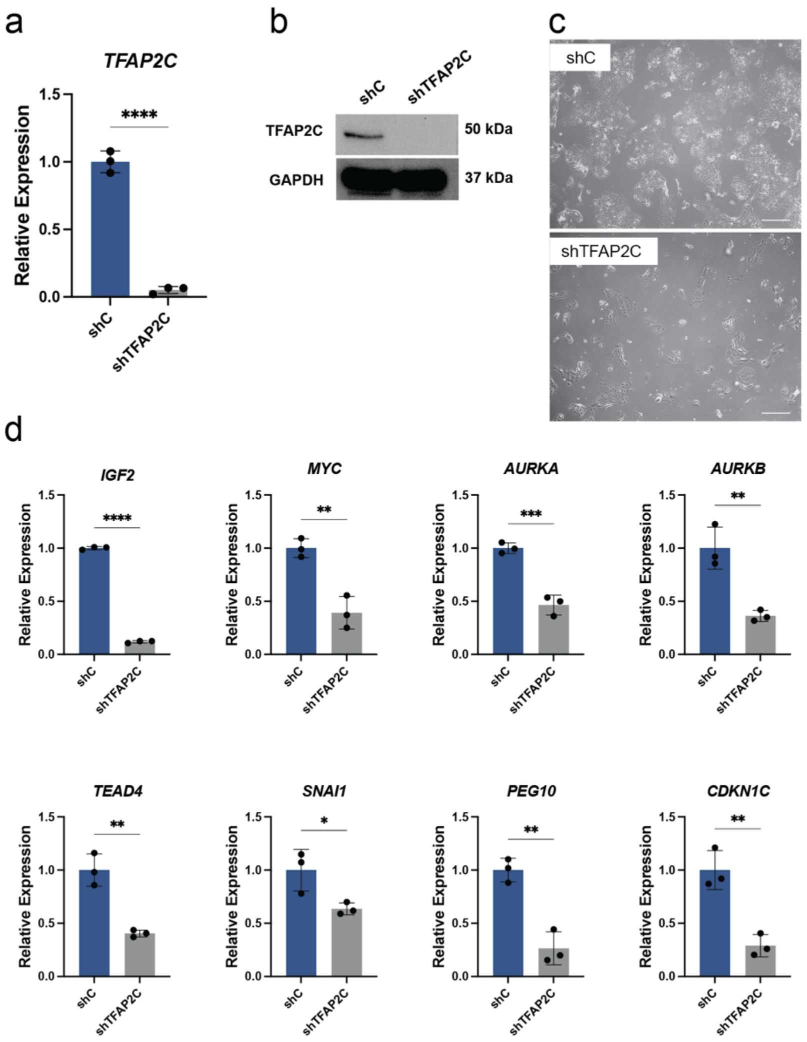

**Supplementary Figure 19. Examination of the impact of trophoblast cell differentiation in CT29 cells upon *TFAP2C* disruption. a)** Relative expression of *TFAP2C* normalized to *POLR2A* in stem state cells transduced with lentivirus containing a control shRNA (**shC**) or a *TFAP2C*-specific shRNA (**shTFAP2C**; n=3 per group; ****p<0.0001). Graph depicts mean ± standard deviation. **b)** TFAP2C (50 kDa) and GAPDH (37 kDa) proteins in stem state cells transduced with shC or shTFAP2C and assessed by western blot. **c)** Phase contrast images of stem state cells transduced with shC or shTFAP2C. Scale bar represents 500 µm. **d)** RT-qPCR measurement of *IGF2*, *MYC*, *AURKA*, *AURKB, TEAD4, SNAI1, PEG10,* and *CDKN1C* in stem state cells transduced with shC or shTFAP2C (n=3 per group) (*p<0.05, **p<0.01, ***p<0.001, ****p<0.0001). Graphs depict mean ± standard deviation.

a

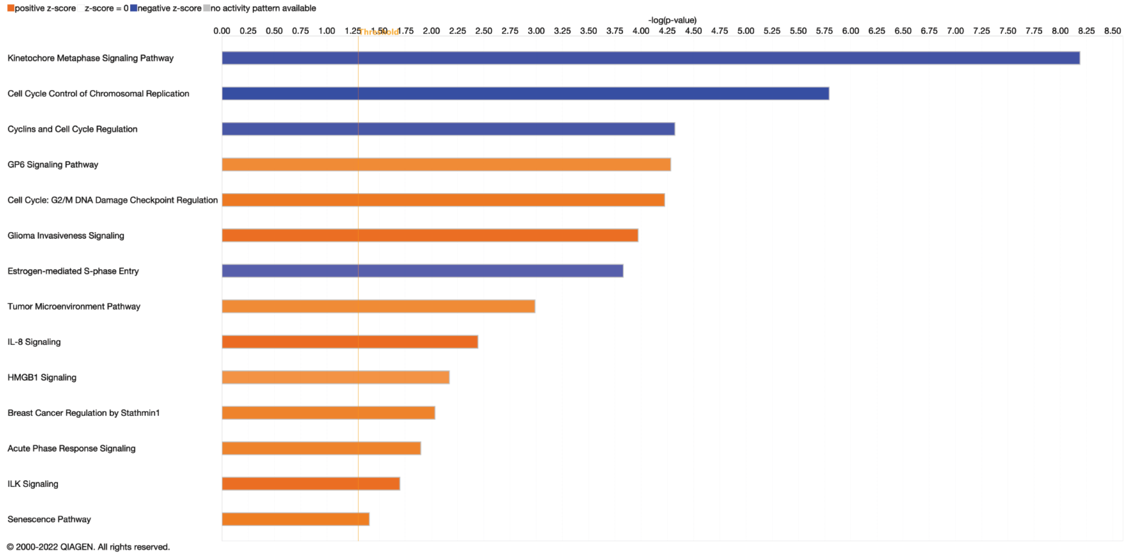

b

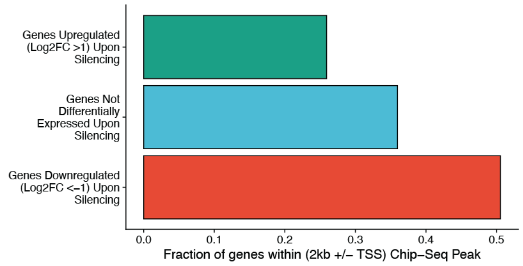

**Supplementary Figure 20. *TFAP2C* regulation of genes linked to cell cycle control. a)** Ingenuity pathway analysis of the top 500 genes regulated by *TFAP2C* (upregulated or downregulated). The top functional pathways (z-score >2) included kinetochore metaphase signaling, cell cycle control of chromosomal replication, as well as cyclins and cell cycle regulation. Positive (orange) and negative (blue) z-scores are presented as histogram bars and -log(B-H p-value) are represented on the x-axis. **b)** Bar graph depicting the fraction of upregulated genes upon *TFAP2C* silencing ($Log_2FC>1$, green bar), not differentially expressed upon *TFAP2C* silencing (blue bar) and downregulated genes upon *TFAP2C* silencing ($Log_2FC<1$, red bar) overlapping a TFAP2C ChIP-Seq peak mapping within 2 kb of the transcription start site (**TSS**) of each gene.

a

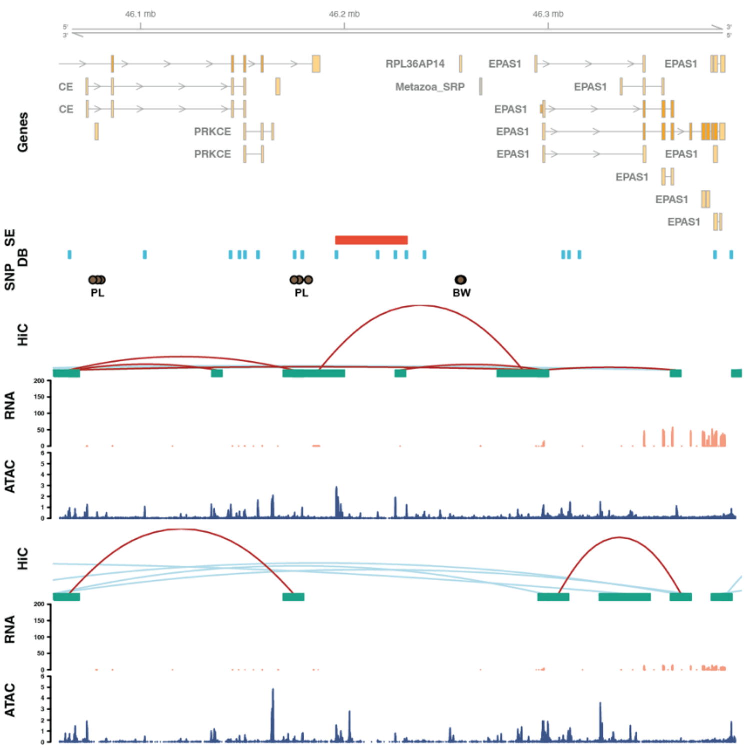

b

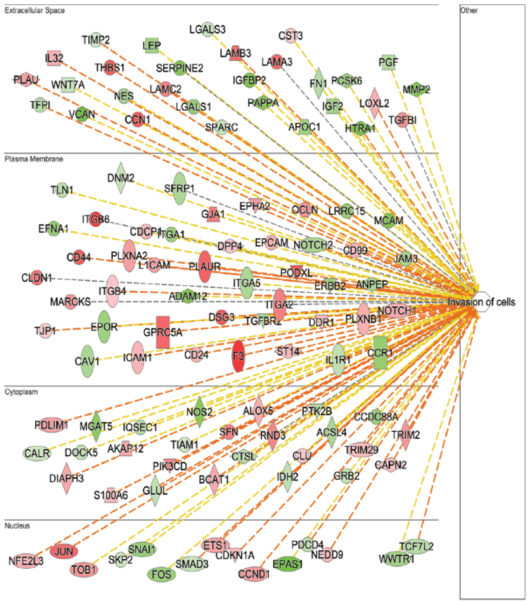

**Supplementary Figure 21. Functional investigation of *EPAS1* on EVT cell differentiation.**

**a)** Hi-C, RNA-Seq, and ATAC-Seq assessments performed in CT29 cells differentiated into EVT cells (top panel) or maintained in the stem state (bottom panel). Regulatory elements near

*EPAS1* are shown and include Hi-C loops (red, both loop anchors in view; blue, loop anchors out of view), RNA-Seq (transcripts per million, **TPM**, y-axis), and ATAC-Seq (counts per million mapped reads, **CPM**, y-axis). All datasets are shown in individual tracks. Super-enhancers (SE, red) and differentially bound regions (**DB**, blue) are specific to the EVT cell state. SNPs associated with pregnancy loss (PL) or birth weight (BW) in large genome-wide association studies (GWAS) are shown in brown circles. **b)** Network schematic depicting results from Ingenuity Pathway Analysis performed on top 500 differentially expressed genes (upregulated or downregulated) by *EPAS1,* in CT27 cells of which 209 genes was annotated to the biological function "Migration of Cells" (p=8.3E-49). The most significant biological function associated with this subset was "Invasion of Cells" (p=5.3E-70). The biological function is displayed as a network with subcellular layout with gene targets as nodes and the node shapes indicate the gene's primary function. Node colors indicate increased (red) or decreased (green) expression of the genes following *EPAS1* disruption.

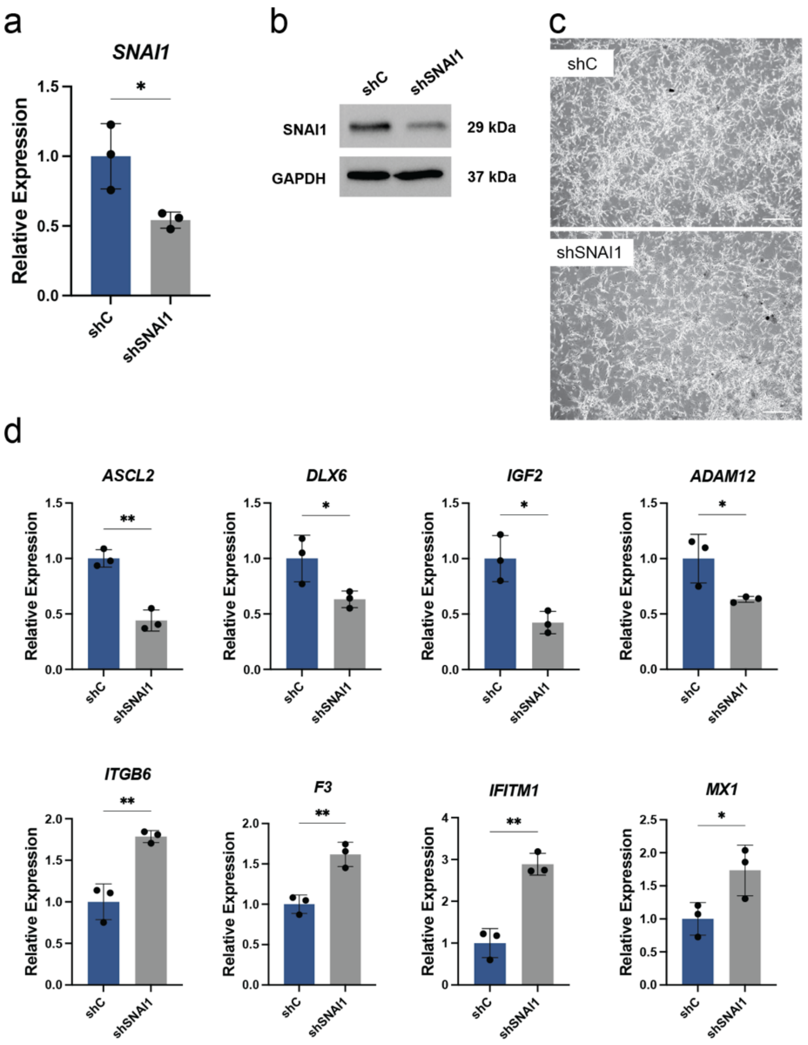

**Supplementary Figure 22.** ***SNAI1*** **disruption impairs EVT cell differentiation in CT29 cells.** **a)** Relative expression of *SNAI1* normalized to *POLR2A* in EVT cells differentiated from stem state cells transduced with lentivirus containing a control shRNA (**shC**) or a *SNAI1*-specific shRNA (**shSNAI1**; n=3 per group; *p<0.05). Graph depicts mean ± standard deviation. **b)** SNAI1 (29 kDa) and GAPDH (37 kDa) proteins in EVT cells differentiated from stem state cells transduced with shC or shSNAI1 and assessed by western blot. **c)** Phase contrast images of EVT cells differentiated from stem state cells transduced with shC or shSNAI1. Scale bar represents 500 µm. **d)** RT-qPCR measurement of *ASCL2*, *DLX6*, *IGF2*, *ADAM12, ITGB6, F3, IFITM1,* and *MX1* in EVT cells differentiated from stem state cells transduced with shC or shSNAI1 (n=3 per group) (*p<0.05, **p<0.01). Graphs depict mean ± standard deviation.

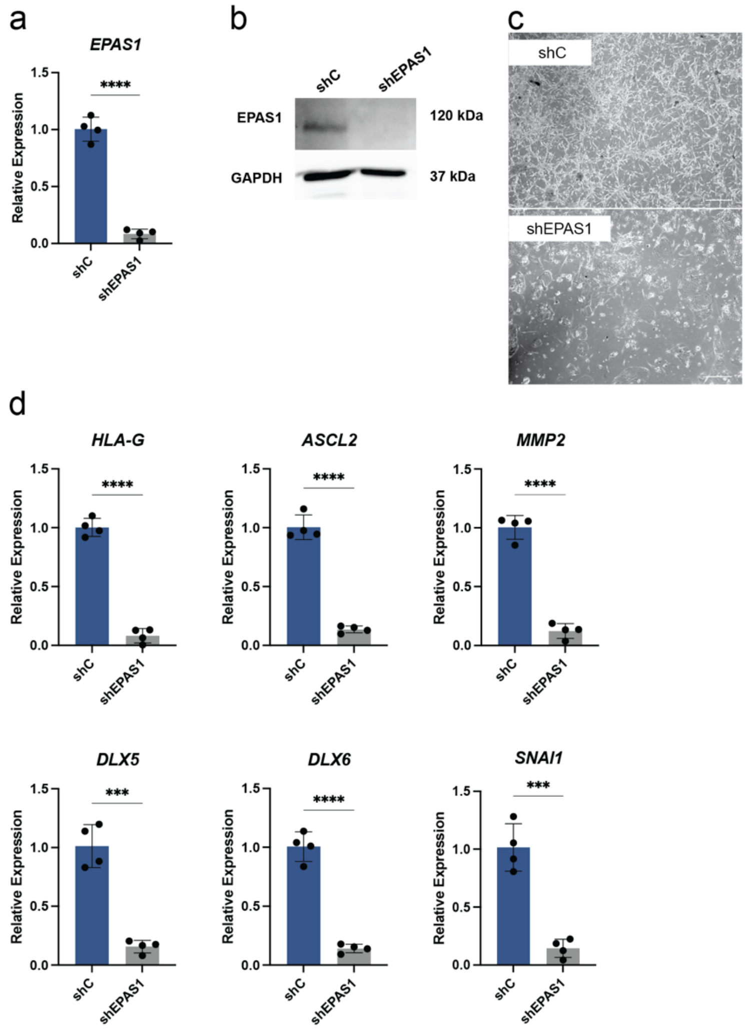

**Supplementary Figure 23.** ***EPAS1*** **disruption impairs EVT cell differentiation in CT29 cells. a)** Relative expression of *EPAS1* normalized to *POLR2A* in EVT cells differentiated from stem state cells transduced with lentivirus containing a control shRNA (**shC**) or a *EPAS1*-specific shRNA (**shEPAS1**; n=3 per group; *p<0.05). Graph depicts mean ± standard deviation. **b)** HIF2a (29 kDa) and GAPDH (37 kDa) proteins in EVT cells differentiated from stem state cells transduced with shC or shEPAS1 and assessed by western blot. **c)** Phase contrast images of EVT cells differentiated from stem state cells transduced with shC or shEPAS1. Scale bar represents 500 µm. **d)** RT-qPCR measurement of *HLA-G*, *ASCL2*, *MMP2*, *DLX5, DLX6,* and *SNAI1* in EVT cells differentiated from stem state cells transduced with shC or shEPAS1 (n=3 per group) (***p<0.001, ****p<0.0001). Graphs depict mean ± standard deviation.

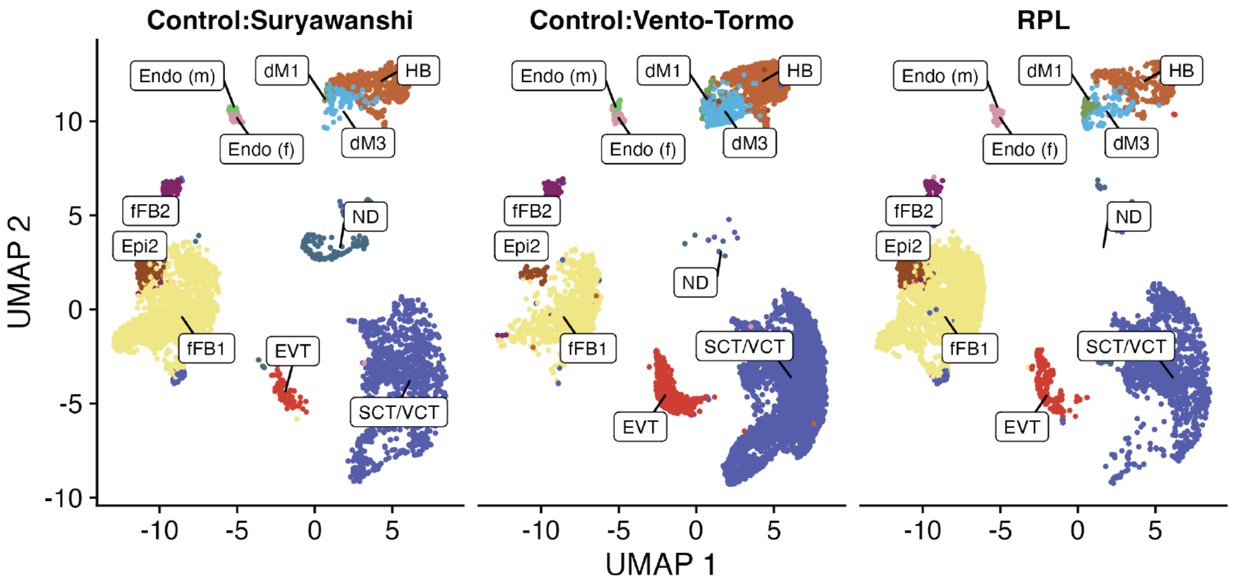

**Supplementary Figure 24. Human placenta gene expression profiles from combined single cell datasets of control and RPL.** UMAP plots depicting cell clustering from two publicly available[2,3] (control; left and middle) and one newly generated (**RPL**; right) single-cell RNA sequencing datasets. Cell types are inferred using marker genes published previously[3] as decidual macrophages (**dM**), fetal (**Endo f**) and maternal endothelial cells (**Endo m**), epithelial glandular cells (**Epi**), extravillous trophoblast cells (**EVT**), fetal fibroblasts (**FB**), Hofbauer cells (**HB**), syncytiotrophoblast and villous cytotrophoblast (**SCT/VCT**) and cells not determined (**ND**).

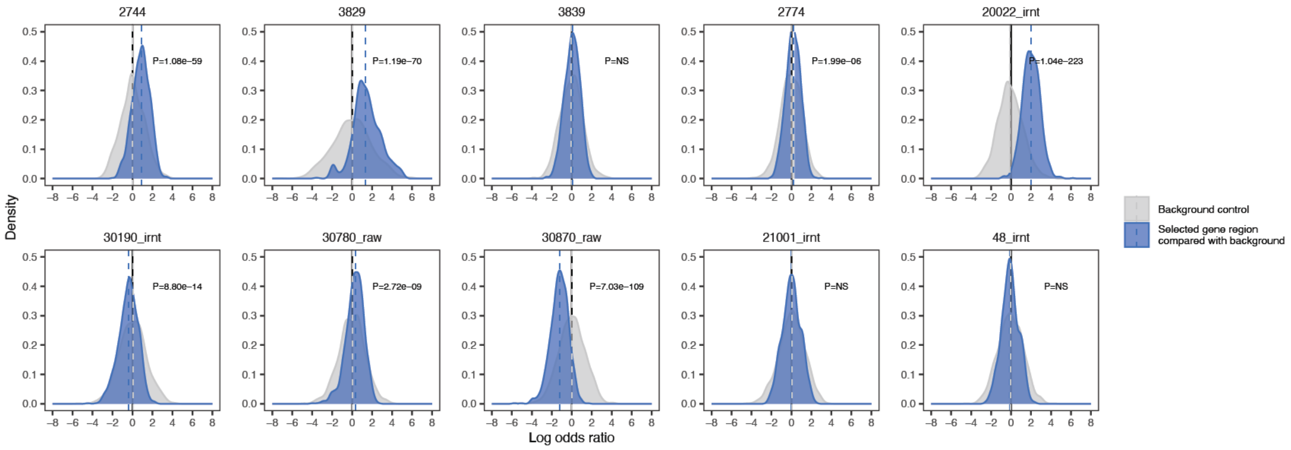

**Supplementary Figure 25. Enrichment analysis of genetic variants near *EPAS1*associated with pregnancy phenotypes in the UK Biobank.** Density plot of the distribution of log transformation of odds ratios (y-axis) for association of genetic variants near *EPAS1* (chr2:46,150,000-46,450,000) and pregnancy phenotypes in UK Biobank compared with genetic variants in randomly selected matched window (blue) with the median odds ratio indicated by dashed blue line. Indicated p-value represent the significance of the odds ratio distribution compared to a null distribution (gray) estimated by the Wilcoxon test. Enrichment analysis was performed on five pregnancy-related phenotypes (2744=Birth weight of first child; 3829=Number of stillbirths; 3839=Number of spontaneous miscarriages; 2774= Ever had stillbirth, spontaneous miscarriage or termination; 20022_irnt=Birth weight) and five randomly selected phenotypes as control (30190_irnt=Monocyte percentage; 30780_raw= LDL direct; 30870_raw=Triglycerides; 21001_irnt= Body mass index (BMI); 48_irnt= Heel bone mineral density (BMD).
